# The 4C Initiative (Clinical Care for Cardiovascular disease in the COVID-19 pandemic) – monitoring the indirect impact of the coronavirus pandemic on services for cardiovascular diseases in the UK

**DOI:** 10.1101/2020.07.10.20151118

**Authors:** 4C Initiative of the CVD-COVID-UK consortium, S Ball, A Banerjee, C Berry, J Boyle, B Bray, W Bradlow, A Chaudhry, R Crawley, J Danesh, A Denniston, F Falter, JD Figueroa, C Hall, H Hemingway, E Jefferson, T Johnson, G King, K Lee, P McKean, SM Mason, N Mills, E Pearson, M Pirmohamed, MTC Poon, R Priedon, A Shah, R Sofat, J Sterne, F Strachan, CLM Sudlow, Z Szarka, W Whiteley, M Wyatt

## Abstract

**Background:** The coronavirus (COVID-19) pandemic affects cardiovascular diseases (CVDs) directly through infection and indirectly through health service reorganisation and public health policy. Real-time data are needed to quantify direct and indirect effects. We aimed to monitor hospital activity for presentation, diagnosis and treatment of CVDs during the pandemic to inform on indirect effects.

**Methods:** We analysed aggregate data on presentations, diagnoses and treatments or procedures for selected CVDs (acute coronary syndromes, heart failure, stroke and transient ischaemic attack, venous thromboembolism, peripheral arterial disease and aortic aneurysm) in UK hospitals before and during the COVID-19 epidemic. We produced an online visualisation tool to enable near real-time monitoring of trends.

**Findings:** Nine hospitals across England and Scotland contributed hospital activity data from 28 Oct 2019 (pre-COVID-19) to 10 May 2020 (pre-easing of lockdown), and for the same weeks during 2018-2019. Across all hospitals, total admissions and emergency department (ED) attendances decreased after lockdown (23 March 2020) by 57.9% (57.1-58.6%) and 52.9% (52.2-53.5%) respectively compared with the previous year. Activity for cardiac, cerebrovascular and other vascular conditions started to decline 1-2 weeks before lockdown, and fell by 31-88% after lockdown, with the greatest reductions observed for coronary artery bypass grafts, carotid endarterectomy, aortic aneurysm repair and peripheral arterial disease procedures. Compared with before the first UK COVID-19 (31 January 2020), activity declined across diseases and specialties between the first case and lockdown (total ED attendances RR 0.94, 0.93-0.95; total hospital admissions RR 0.96, 0.95-0.97) and after lockdown (attendances RR 0.63, 0.62-0.64; admissions RR 0.59, 0.57-0.60). There was limited recovery towards usual levels of some activities from mid-April 2020.

**Interpretation:** Substantial reductions in total and cardiovascular activities are likely to contribute to a major burden of indirect effects of the pandemic, suggesting they should be monitored and mitigated urgently.

**Funding:** British Heart Foundation, Health Data Research UK

## Background

Beyond direct effects of the coronavirus disease 2019 (COVID-19) on individuals and populations in every country, the pandemic has had and will continue to have indirect effects on morbidity and mortality, through changes in patient and clinician behaviour, and health system reorganisation and/or strain.^1^ In order to plan and adapt responses to this and future public health threats, therefore, there is a need to understand the indirect effects of the pandemic on non-COVID diseases, particularly non-communicable diseases (NCDs), for both disease burden and health service provision.^2^

Cardiovascular diseases (CVD) are the largest cause of morbidity and mortality in the UK and globally.^3,4^ Moreover, prior CVD is a major risk factor for complications and mortality associated with COVID-19.^5,6^ Government guidance has advised individuals with CVD to pay particular attention to physical isolation measures.^7^ Concerns have been raised about provision of care during the pandemic for these diseases across the spectrum, from prevention to treatment, and are supported by data from multiple countries showing reduced service level activity.^8–10^ In the UK, studies have reported reductions in activity across CVDs.^11,12^ Moreover, official national statistics show an excess of non-COVID and CVD deaths,^13,14^ as well as a reduction in emergency department (ED) attendances for cardiac presentations.^15^

There are multiple disease-specific national audits for CVD, in addition to routine primary and secondary care data. However, data from these sources often lag several weeks or even months behind real time, may not include the UK’s devolved nations (Scotland, Wales and Northern Ireland) and are not currently widely accessible for analyses.^16^ For audit, quality improvement, surveillance and to inform policy responses, it is necessary to determine changes in service delivery which have occurred during the pandemic at hospital and national levels. Transparent, public-facing, near real-time information has been shown to be of value to patients, the public, researchers, clinicians and policymakers alike during the pandemic.^1,17^

### Objectives

For presentation, diagnosis and treatment of CVD, we aimed to: (i) develop a protocol for national surveillance of CVD hospital services during the COVID-19 pandemic; (ii) present pilot data from a preliminary cohort of hospitals; and (iii) design and implement a simple tool for monitoring and visualising trends in CVD hospital services in the UK.

## Methods

### Study design and data sources

We conducted a retrospective hospital-based analysis of presentations, diagnoses and treatments or procedures for selected CVDs in hospitals across the UK before and during the COVID-19 epidemic. The protocol was developed by a group of seven cardiovascular clinicians with relevant clinical, epidemiological and health data science expertise and agreed with members of the CVD-COVID-UK collaboration, supported by the BHF Data Science Centre.^18^

### Data collection

We sent email invitations to contribute data to the Chief Clinical Information Officers or other relevant contacts in 21 hospitals (or groups of regionally connected hospitals) across the UK. We requested aggregate data, with no individual level or linked data, including inpatient admissions and ED visits, both overall and for specified cardiovascular diagnoses and procedures or treatments, and we provided guidance on which ICD-10, OPCS-4 or equivalent codes to use. We selected six disease areas to gain an overview of CVD service provision in a short timescale:

- Acute coronary syndromes (ACS)
- Heart failure (HF)
- Stroke and transient ischaemic attack (TIA)
- Venous thromboembolism (VTE)
- Peripheral arterial disease (PAD)
- Aortic aneurysm

Supplementary Appendix 1 shows the data request sent to hospitals for weekly counts of relevant ED categories, hospital admission diagnoses and hospital treatments/procedures covering the period 28 October 2019 to 10 May 2020, designed to show activity in three phases: pre-COVID-19 pandemic to the first UK confirmed case of COVID-19 (31 January 2020), between the first COVID-19 case but pre-lockdown on 23 March 2020, and post-lockdown.

Since our principal aim was to capture change in activity over time within each hospital rather than to make direct comparisons between hospitals, we incorporated a degree of flexibility in the way each data item was defined. For example, one hospital which did not have timely access to ICD codes for ACS was able to provide counts using a data capture tool embedded into the patient record that identifies all patients with ACS at the point of cardiac troponin testing.^19^ Where available, as measures of total acute activity, we requested counts of all ED attendances, all hospital admissions and all COVID-19 hospital admissions.

In addition, given known seasonal fluctuations in hospital services, we requested data on the same items from the same calendar weeks (or months) in the previous year (2018-2019) for comparison. We recognised that not all disease areas or treatments and procedures would be relevant to or available from every participating hospital, and so invited hospitals to contribute whichever data items they were able to.

### Data analysis

The study period was divided into three phases based on complete weeks (Monday-Sunday) of data: (i) before the first case of SARS CoV2 in the UK (28 Oct 2019 to 2 Feb 2020 [14 weeks]); (ii) between the first case and start of lockdown in the UK (3 Feb 2020 to 22 Mar 2020 [7 weeks]): and (iii) during lockdown (23 Mar 2020 to 10 May 2020 [7 weeks]).

We used the aggregate data to compare the weekly counts within each hospital and across all hospitals combined, estimating associated 95% confidence intervals (CIs) using the Poisson exact method. We calculated means of hospital statistics and their 95% CIs in the combined analyses. In addition, we calculated percentage change from the corresponding week in the previous year (2018-2019), together with 95% CIs using the Wilson score interval, in the three phases described above. We also calculated the percentage reduction in total hospital admissions and ED attendances around Christmas and New Year in 2019-2020 by comparing the period 16 December 2019 to 12 January 2020 to the immediately preceding period 18 November to 15 December 2019, using the same approach with equivalent weeks to calculate the percentage reduction around Christmas and New Year in 2018-2019. We calculated 4-week rolling means, that is, the means of hospital activities over each 4-week period, starting 3 weeks prior to the week of interest. Such rolling means enable clearer visualisation of trends for data items with low numbers and high variability. Where data providers suppressed values less than 5, we converted these to the value of 2.5 for analytic purposes. To demonstrate relative change of hospital activities between phases, we also calculated relative reduction in weekly counts compared with the first phase (before the first UK case of COVID-19). We performed all statistical procedures in R (version 4.0.0). Relevant code is available at https://github.com/HDRUK/4C-Initiative.

### Open access interactive data visualisation tool

As understanding trends and patterns in hospital activity should inform policymakers and other stakeholders in the planning of services as the UK eases out of lockdown, we developed an interactive online tool to enable dynamic visualisation of the data. It will enable periodic (e.g., monthly) updates of data from contributing hospitals to monitor trends over time and facilitate incorporation of data from other UK hospitals and comparisons between hospitals (http://www.hospitalactivity.com).

## Results

### Overall hospital activity

Of 21 hospitals (or groups of regionally connected hospitals) contacted, 17 agreed to participate. Of these, nine hospitals distributed across England and Scotland, providing cardiovascular services to an estimated population of up to 10 million people, contributed data in time for the analyses reported here. There were 513,703 hospital admissions from eight hospitals and 435,653 ED attendances from five hospitals during the period 28 Oct 2019 to 10 May 2020 compared with 599,372 and 506,516 respectively in 2018-2019. There were 676 and 5,182 COVID-related admissions from eight hospitals for the second and third phases (after the first UK case of COVID until lockdown commenced, and from the start of lockdown onwards) respectively.

Across all hospitals, total admissions before the first case of COVID-19 were very similar to the corresponding period in 2018-2019, including the expected dip in admissions of around 12% during the Christmas period in December 2018 and 2019 (Supplementary Table 1). After lockdown, admissions decreased by 57.9% (95% CI 57.1-58.6%) compared with the previous year (Table 1 and Figure 1).

**Table 1.** Hospital statistics as percentage change from corresponding dates in 2018-2019

|  |  | Percentage change from 2018-2019 |  |  |  |  |  |
| --- | --- | --- | --- | --- | --- | --- | --- |
|  | No. of hospitals | Before 1<sup>st</sup> case |  | Between 1<sup>st</sup> case and lockdown |  | After lockdown |  |
|  |  | % | 95% CI | % | 95% CI | % | 95% CI |
| **Overall** |  |  |  |  |  |  |  |
| Total ED attendances | 5 | 3.4 | 3.2 to 3.6 | -8.8 | -8.4 to -9.1 | -52.8 | -52.2 to -53.5 |
| Total hospital admissions | 8 | 1.1 | 1.0 to 1.2 | -6.3 | -6.0 to -6.7 | -58.2 | -57.5 to -58.9 |
| **Cardiac** |  |  |  |  |  |  |  |
| ED attendance with cardiac conditions | 4 | 5.7 | 4.3 to 7.6 | -9.6 | -7.2 to -12.8 | -40.2 | -35.6 to -45.0 |
| Admission with ACS | 9 | -1.7 | -1.1 to -2.6 | -15.7 | -13.0 to -18.9 | -39.4 | -35.3 to -43.5 |
| Admission with heart failure | 7 | 6.1 | 5.1 to 7.3 | -3.2 | -2.2 to -4.5 | -49.0 | -45.7 to -52.2 |
| PCI performed | 7 | -6.9 | -5.0 to -9.4 | -8.2 | -5.4 to -12.2 | -39.6 | -33.7 to -45.8 |
| Cardiac pacemaker and resynchronisation | 8 | 2.3 | 1.0 to 4.9 | 0.0 | 0.0 to 2.8 | -47.2 | -38.8 to -55.9 |
| CABG performed | 6 | -9.4 | -5.0 to -16.9 | -9.8 | -4.3 to -21.0 | -69.6 | -55.2 to -80.9 |
| **Cerebrovascular** |  |  |  |  |  |  |  |
| ED attendance with cerebrovascular conditions | 4 | -1.9 | -1.0 to -3.5 | -6.5 | -4.0 to -10.2 | -31.8 | -26.2 to -38.0 |
| Admission with acute stroke/TIA | 6 | -7.5 | -5.8 to -9.8 | -11.9 | -8.8 to -15.8 | -49.2 | -43.7 to -54.7 |
| Stroke thrombolysis and thrombectomy | 5 | -5.6 | -1.0 to -25.8 | 0.0 | 0.0 to 25.9 | -45.5 | -21.3 to -72.0 |
| Carotid endarterectomy / stenting | 4 | 30.8 | 12.7 to 57.6 | 25.0 | 7.1 to 59.1 | -66.7 | -30.0 to -90.3 |
| Cerebral aneurysm coiling | 5 | -9.6 | -5.7 to -15.7 | -35.8 | -26.9 to -45.8 | -59.4 | -47.1 to -70.5 |
| **Other vascular** |  |  |  |  |  |  |  |
| ED attendance with vascular conditions | 3 | 0.6 | 0.1 to 3.2 | -16.0 | -9.9 to -24.7 | -40.6 | -31.5 to -50.3 |
| Admission with aortic aneurysms | 7 | 13.7 | 10.1 to 18.2 | 9.4 | 5.5 to 15.3 | -53.0 | -44.5 to -61.3 |
| Admission with peripheral arterial disease | 6 | 14.4 | 12.4 to 16.8 | 2.8 | 1.7 to 4.7 | -49.2 | -44.8 to -53.6 |
| Admission with DVT or PE | 6 | 11.5 | 8.6 to 15.0 | -12.9 | -8.9 to -18.2 | -37.2 | -30.6 to -44.2 |
| Limb revascularisation, bypass or amputation | 6 | -1.2 | -0.4 to -3.3 | -3.7 | -1.6 to -8.3 | -68.2 | -59.8 to -75.5 |
| Aortic aneurysm repair | 6 | -18.8 | -10.2 to -31.9 | -20.8 | -9.2 to -40.5 | -88.2 | -65.7 to -96.7 |
| Peripheral angioplasty | 6 | 15.0 | 10.1 to 21.6 | 9.1 | 4.5 to 17.6 | -65.5 | -54.8 to -74.8 |

ED = accident & emergency; ACS = acute coronary syndrome; PCI = percutaneous coronary intervention; CABG = coronary artery bypass graft; TIA = transient ischaemic attack; DVT = deep vein thrombosis; PE = pulmonary embolism; CI = confidence interval; before 1<sup>st</sup> case = 28 Oct 19 to 2 Feb 20; between 1<sup>st</sup> case and lockdown = 3 Feb 20 to 22 Mar 20; after lockdown = 23 Mar 20 to 10 May 20.

**Figure 1.**
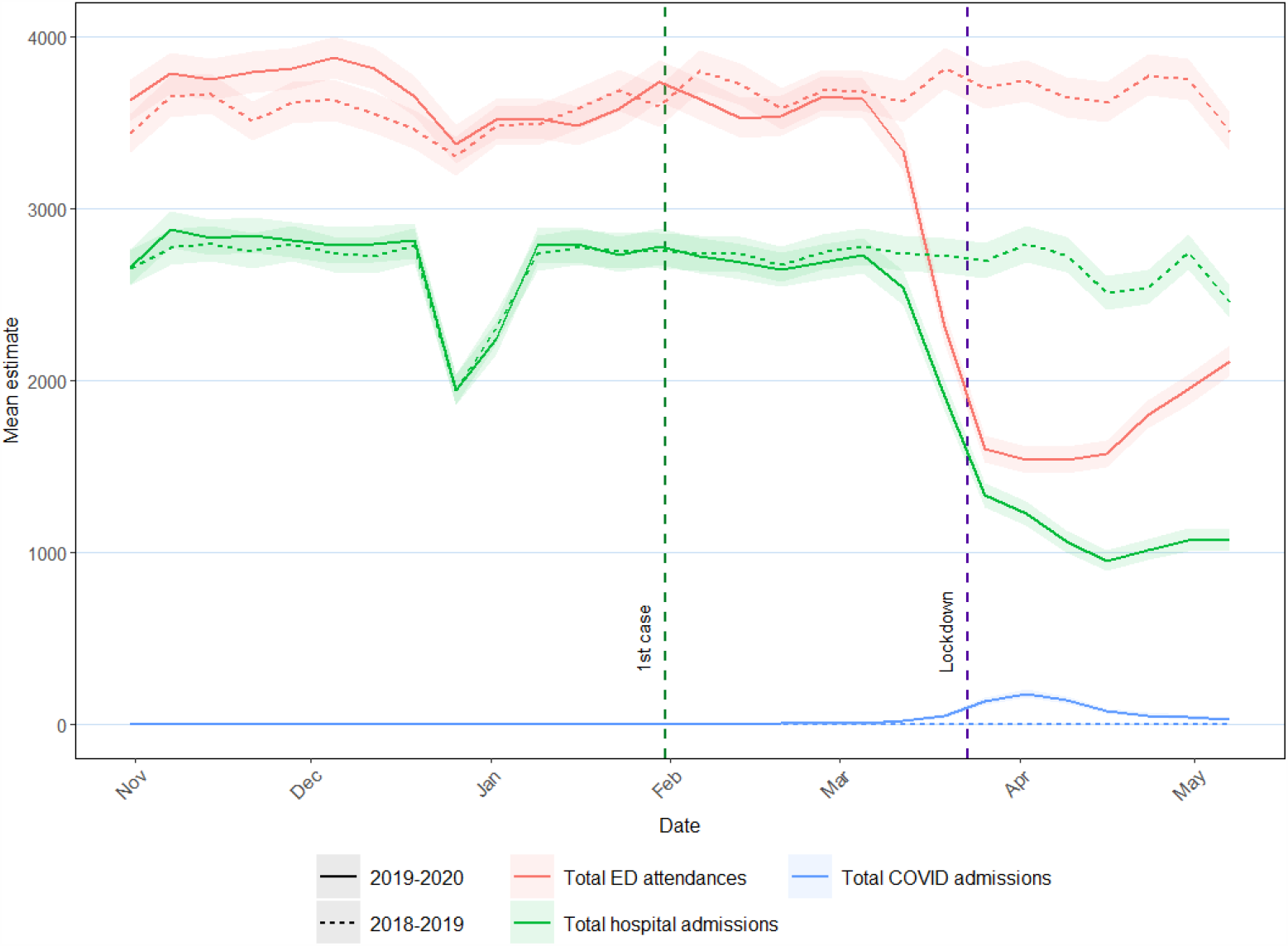
Overall hospital activity (admissions, ED attendances and COVID admissions) between 31 Oct 2019 and 10 May 2020 compared with the same weeks from 2018-2019. Lines describe the mean hospital activities in 2019-2020 (solid) and 2018-2019 (dotted). Shading represents 95% confidence interval of the respective hospital activity. The first case of COVID-19 was on 31 January 2020 and lockdown started on 23 March 2020. ED = Emergency Department.

Overall ED attendances showed a similar pattern, with a small 3.4% (3.2-3.5%) increase compared with the previous year before the first case, a more modest winter reduction of around 8% (Supplementary Table 1), a marked 52.9% (52.2-53.5%) reduction after lockdown and evidence of some recovery back towards pre-COVID levels of activity from mid-April 2020. The reduction in hospital admissions was substantially larger than the number of admissions for COVID-19 (Figure 1). Overall hospital admission and ED attendance patterns were generally consistent across individual hospitals. However, while there was some recovery in ED attendances from mid-April 2020 in all hospitals that provided data, hospital admissions had only started to return towards usual levels in some, but not all, hospitals by early May (Figure 2).

**Figure 2.**
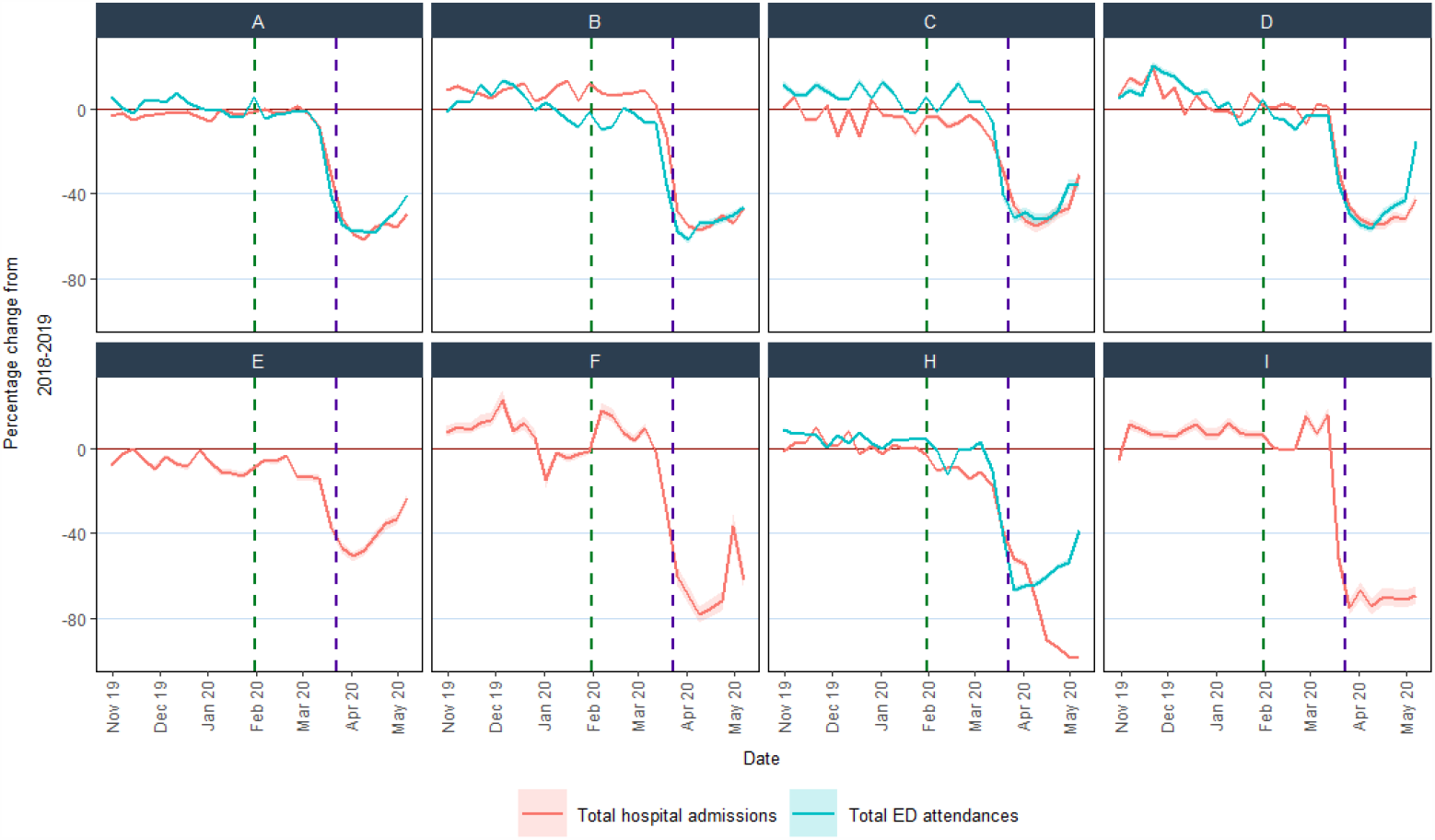
% change compared with the previous year in ED attendances and hospital admissions for individual hospitals. Eight hospitals provided data on hospital admissions and five hospitals (A, B, C, D, and H) also provided data on ED attendances. Hospital G did not provide these hospital statistics and is not shown.

### Cardiac, cerebrovascular and other vascular conditions

Compared with the previous year, hospital statistics on cardiac, cerebrovascular and other vascular conditions dropped by between 31% and 88% after lockdown (Table 1). Most started to decline 1-2 weeks before the lockdown. Some recovery from mid-April 2020 was evident in ED attendances for these conditions and for cardiac procedures (primarily driven by percutaneous coronary intervention) (Figure 2 and www.hospitalactivity.com). The greatest proportional reductions (65% or more) were in coronary artery bypass graft surgery, carotid endarterectomy, aortic aneurysm repair and procedures for peripheral arterial disease (Table 1).

### Comparison with the pre-COVID period

Compared with the period before the first COVID-19 case, activity declined across diseases and specialties between the first case and lockdown (total ED attendances RR 0.94, 0.93-0.95; total hospital admissions RR 0.96, 0.95-0.97) and, more markedly, after lockdown (total ED attendances RR 0.63, 0.62-0.64; total hospital admissions RR 0.59, 0.57-0.60). Reductions in coronary artery bypass grafts, carotid endarterectomy, peripheral arterial procedures and aortic aneurysm repairs were the most prominent (RRs 0.23 to 0.49). (Figure 4). All hospitals had reduced activities after lockdown but with variation between hospitals in the extent of reduction for different diseases and treatments (Supplementary Figure 1).

**Figure 3.**
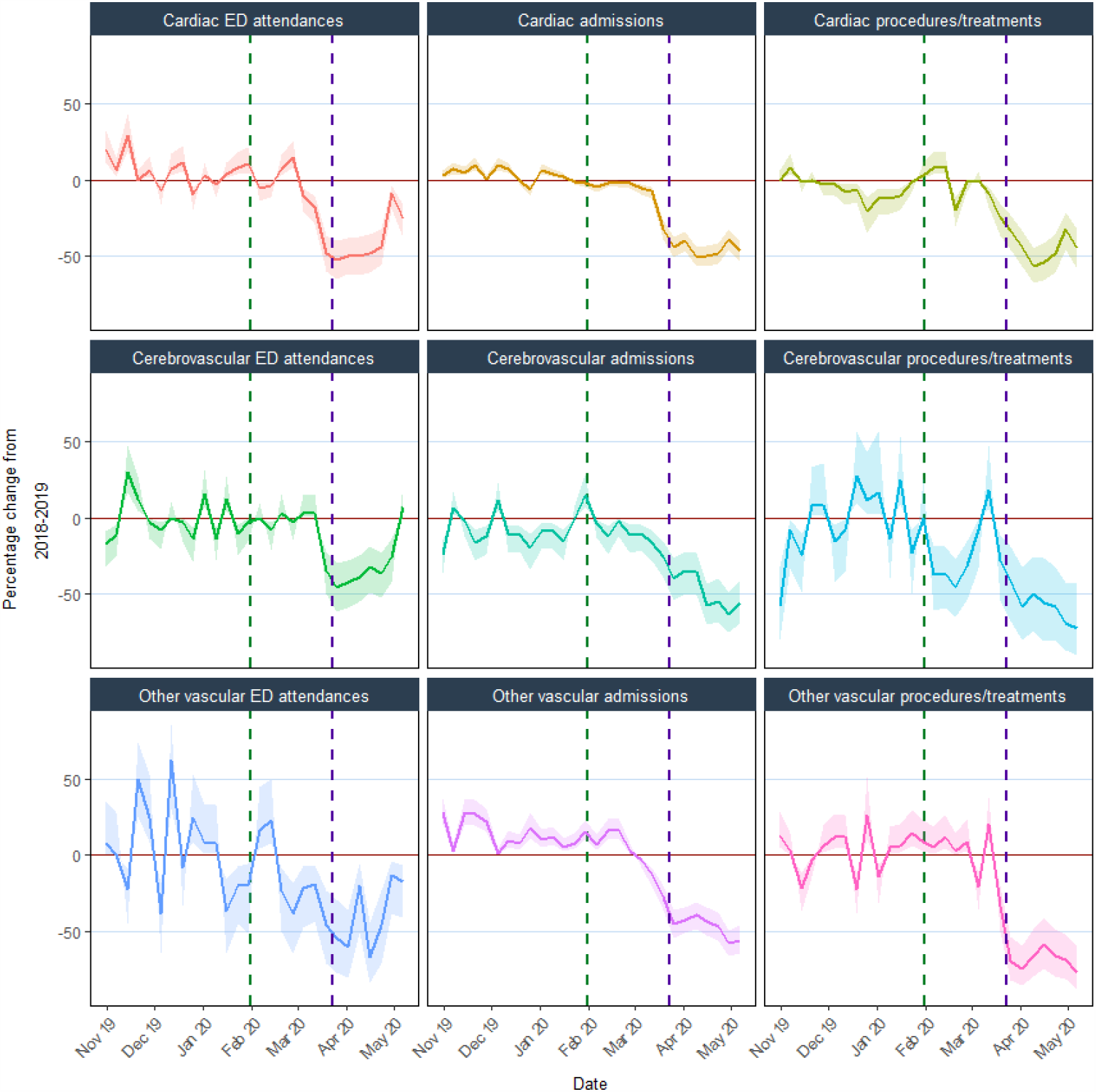
% change compared with the previous year in ED attendance, hospital admissions, and procedures/treatments for cardiac, cerebrovascular and other vascular conditions. Cardiac ED attendances are those with an ED diagnosis code for cardiac conditions; cardiac admissions include those with acute coronary syndrome or heart failure; cardiac procedures/treatments include percutaneous coronary intervention, cardiac pacemaker or resynchronisation, and coronary artery bypass graft; cerebrovascular ED attendances are those with an ED diagnosis code for cerebrovascular conditions; cerebrovascular admissions include those with acute stroke (ischaemic, intracerebral haemorrhage or subarachnoid haemorrhage) or transient ischaemic attack; cerebrovascular procedures/treatments include stroke thrombolysis, thrombectomy, carotid endarterectomy/stenting, or cerebral aneurysm coiling; other vascular ED attendances are those with an ED diagnosis code for other vascular conditions; other vascular admissions include those with aortic aneurysms, DVT, PE, or peripheral arterial disease; other vascular procedures include aortic aneurysm repair, limb revascularisation, bypass or amputation, and peripheral angioplasty. Horizontal brown line indicates 0%; vertical green dotted line indicates first confirmed COVID-19 case on 31 Jan 20; vertical purple dotted line indicates lockdown date on 23 Mar 20. Shading represents 95% CIs of % change.

**Figure 4.**
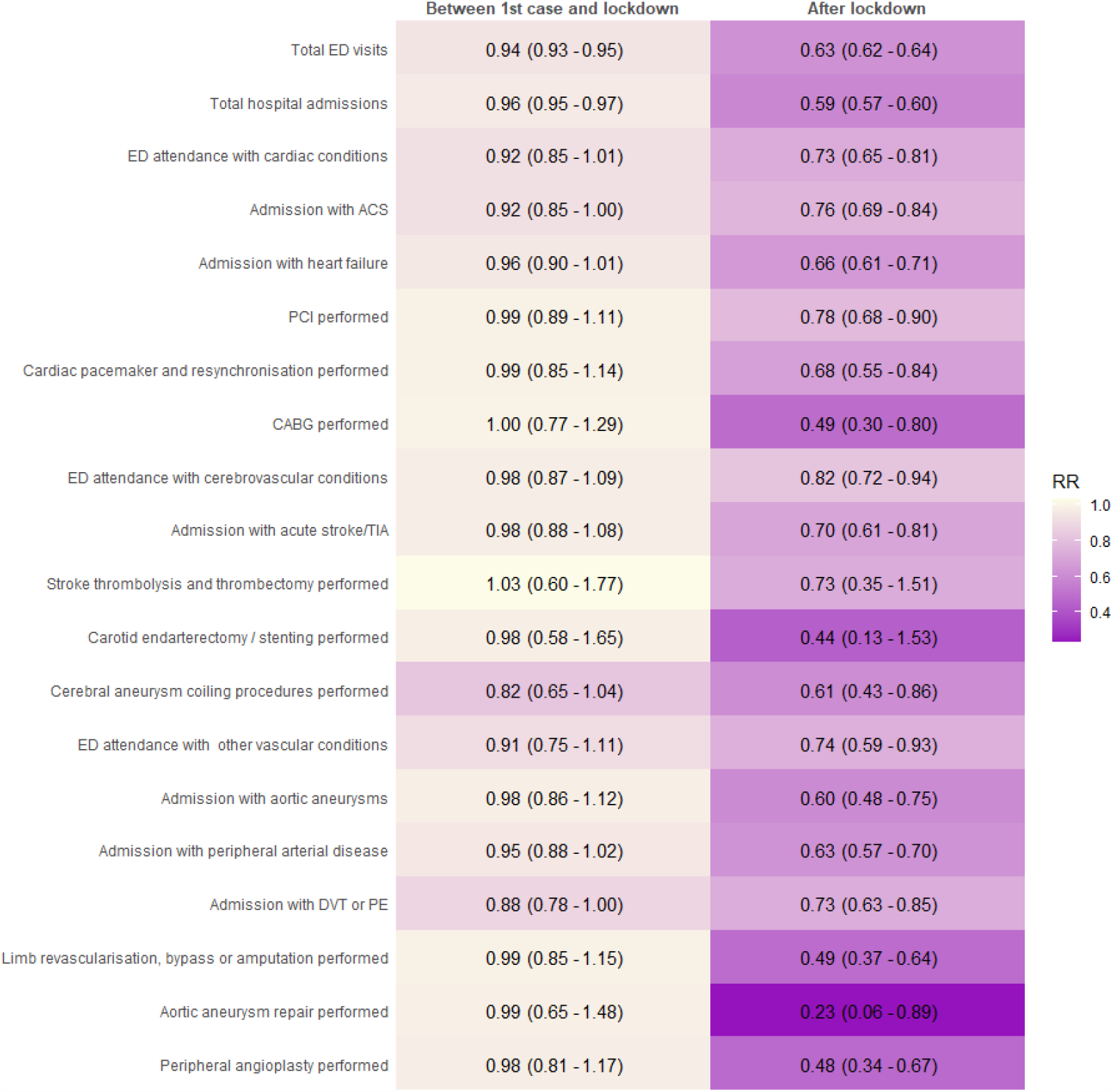
Relative reductions in hospital activities during the COVID-19 pandemic. Relative risks (RR) comparing phase 2 (between first case and lockdown) and phase 3 (after lockdown) to phase 1 (before first case). ED: emergency department; ACS: acute coronary syndromes; PCI: percutaneous coronary interventions; CABG: coronary artery bypass graft; TIA: transient ischaemic attack; DVT: deep vein thrombosis; PE: pulmonary embolus.

## Discussion

### Summary of findings

Three main findings have emerged from this rapid assimilation of UK hospital routine electronic health record data. First, the current data have shown in detail the pattern of sharp reductions in activity across CVDs, across services and across hospitals, during the initial wave of the pandemic. Second, the results have demonstrated that changes in hospital services started to occur prior to lockdown, with some – but not all – moving towards pre-COVID activity levels by early May 2020. Third, the visualisation tool developed for this analysis has illustrated how aggregate data can be collected from hospitals and presented rapidly.

The current results indicate that the reduction in overall admissions was substantially larger than the increase in COVID-related admissions across all hospitals that provided information on both. Adapting hospital services to address the pandemic has been far more complex than simply accommodating an increase in hospital admissions. Patients with COVID-19 are very resource intensive, with a substantial proportion requiring high dependency or intensive care. Hence, hospitals have had to create additional critical care capacity, through re-purposing both staff and wards. In particular, the need to re-purpose operating theatre staff and operating theatres has required the cancellation of many elective surgical procedures. Hospital-wide measures have also been required to minimise spread of infection. All of these service adaptations have necessitated reductions in overall activity. At the same time, primary care practice in the UK shifted to a total triage system and remote consultation, wherever possible, with unknown consequences, including impact on referral to hospitals. All of these service adaptations were justifiable, but the marked reduction in overall hospital admissions compared with the numbers due to COVID-19 raises the possibility of overall health service over-compensation. The observed reduction in hospital activity will undoubtedly lead to adverse, indirect, long-term impacts on care (which we measure here) and incidence of a wide range of non-COVID diseases (which we do not measure). Understanding the patient and professional behaviours and health service organisational factors contributing to the observed response across different parts of the health service should help in planning appropriate service adaptation to deal with any potential further surge of COVID-19 or future epidemic and pandemic emergencies.

The present data also showed a dramatic and consistent decline in overall admissions and ED attendances from around 2 weeks before lockdown. This may reflect changes in behaviour of clinicians, hospital management, public health and the public, which were occurring prior to lockdown. There is the possibility that disease incidence of both cardiovascular and non-cardiovascular conditions decreased, which needs to be investigated. There was evidence of recovery in ED attendances and in admissions in some hospitals from around mid-April 2020, although, as of 10 May 2020, activity generally remained well below that of the previous year. We also observed marked reductions in CVD-specific ED attendances, admissions and hospital procedures and treatments, albeit with some recovery in ED attendances and percutaneous coronary interventions from mid-April 2020.

We assessed relative change in activity both through percentage change compared with the previous year and through relative reduction compared with before the first UK COVID-19 case. These two approaches yielded consistent results, suggesting that the observed reductions related mainly to the COVID-19 pandemic rather than to year on year trends (for example, the known declining rates in carotid endarterectomy^20^or seasonal fluctuations). Reductions in cardiovascular surgical procedures and procedures for peripheral arterial disease were particularly dramatic, with no evidence of recovery by early May. The likelihood of coronary artery bypass surgery was more than halved during the pandemic with a less marked reduction in percutaneous coronary interventions (Figure 4 and Table 1). Whilst we did not collect data to fully explore the reasons for this disparity between modes of coronary revascularisation, several factors may be relevant. The repurposing of cardiac surgical and anaesthetic resources during the pandemic will underpin the reduction in cardiac surgical procedures. Furthermore, some patients who had been referred or accepted for bypass surgery were redirected in some hospitals to be treated instead by percutaneous coronary intervention which does not require anaesthetic support. Guidelines from the UK’s Vascular Society early in the pandemic suggested increasing the size threshold for elective surgical intervention for abdominal aortic aneurysms and the avoidance of carotid endarterectomy^21^. This advice will undoubtedly have contributed to the dramatic decline in aortic aneurysm repairs and carotid endarterectomies. These disease- and service-specific empirical data enable monitoring of service activity as well as estimates to inform on the indirect effects of the pandemic on morbidity, mortality and health economic measures.^1,11^

We have demonstrated that hospitals across the UK were willing and able to rapidly provide aggregate data to monitor trends in overall and cardiovascular specific activity in close to real-time. Further, we have developed an online tool to facilitate the inclusion of additional hospitals and incorporate data updates for ongoing monitoring of trends in hospital activity as lockdown restrictions ease across the UK in the coming weeks and months. These data and our tool provide surveillance of overall and cardiovascular hospital activities and could inform which services for which diseases require particular attention at system level and at hospital level. This framework could be used in other non-COVID diseases, e.g. cancer and respiratory disease, where national efforts are already underway (e.g. those led by the Health Data Research UK cancer and respiratory research hubs, DATACAN and BREATHE, respectively).^22,23^ These types of data need to be integrated across disease-specific domains in order to tackle the complex nature of the indirect effects of the pandemic as well as the prominent role of multi-morbidity in the risk of COVID-19 severity and mortality. Such data are not currently part of routine pandemic or emergency preparedness^24^ but the scale of the indirect effects across the UK and worldwide suggests that this situation needs to change.

### Comparison with other data

National mortality data from the UK and other countries also demonstrate the direct and indirect impacts of the pandemic, showing peaks in COVID deaths, non-COVID deaths and CVD deaths.^11,25,26^ Other studies have also reported reductions in hospital activity overall, for CVDs and for other conditions (e.g., cancer) during the COVID-19 pandemic, both in the UK and many other countries.^9,27–29^ Where reported, recovery of activity has generally been slow; for example, in hospitals in China, cardiovascular disease activity remained below pre-COVID-19 levels for two to three months, even after easing lockdown.^11^ To our knowledge, the present report is the first UK study to have recorded overall and cardiovascular hospital activity over a long enough period since lockdown to show the beginning of recovery in some measures, and is the only study we are aware of to provide analyses of these via an online tool designed to include data from additional hospitals and regular updates over the months ahead.

### Strengths and limitations

The simple, aggregate nature of our data request enabled a large proportion of the hospitals contacted to provide data in a short timeframe. This has made possible the notion of a regularly updated online tool that can incorporate and display near real-time data from an increasing number of hospitals across the UK. However, the aggregate nature of the data means that the influence of individual level factors such as age, socioeconomic status, ethnicity and comorbidities cannot be explored. Further, our current data request combines data on the primary and other (secondary) reasons for hospital admission, and does not subdivide admissions or procedures according to elective (planned) and emergency (unplanned) activity. Future modifications of our data collection procedures could enable separate analysis of elective and emergency procedures. Finally, we used data from the previous calendar year as a comparator to calculate percentage change in activity. While data averaged across the previous five years may provide a more stable comparator and has been used to assess excess mortality from national mortality data, such data would mask longer term trends in some activities (e.g. reductions in carotid endarterectomies and aortic aneurysm repairs and increasing numbers of procedures for peripheral arterial disease^30^), hospitals may have found it more challenging to provide these data, and changes in hospital catchment areas and service arrangements would be more likely to have occurred over a longer period.

## Conclusion

In conclusion, we have shown the value of simple aggregate data for monitoring changes in general and disease-specific hospital activity during the course of the COVID-19 pandemic in the UK, and the potential for further development of an online tool to enable ongoing monitoring. This will enable individual hospitals to compare activity in their hospital with others, and could provide real-time data to inform the planning and prioritisation of service responses to the current and future public health emergencies.

## Data Availability

Visualisation of data is available in the web application. Raw data is not released because of data governance.

http://hospitalactivity.com

https://github.com/HDRUK/4C-Initiative

## Acknowledgements

We thank staff in the Golden Jubilee National Hospital, Glasgow, including Lynn Hay, Jim Christie and Brian Lawson.

---

**Supplementary Table 1.** Percentage reduction in hospital admissions and ED attendances during the Christmas and New Year period*.

|  | **% reduction 2018-2019**<br>**(95% CI)** | **% reduction 2019-2020**<br>**(95% CI)** | **% reduction pooled across**<br>**both years (95% CI)** |
| --- | --- | --- | --- |
| Hospital admissions | 11.2 (10.7-11.8) | 13.0 (12.4-13.6) | 12.1 (11.7-12.6) |
| ED attendances | 4.0 (3.7-4.4) | 8.0 (7.6-8.4) | 6.1 (5.8-6.4) |

\* comparing the period 16 December 2019 to 12 January 2020 to the immediately preceding period 18 November to 15 December 2019 (and equivalent weeks for the previous year)

**Supplementary Figure 1.**
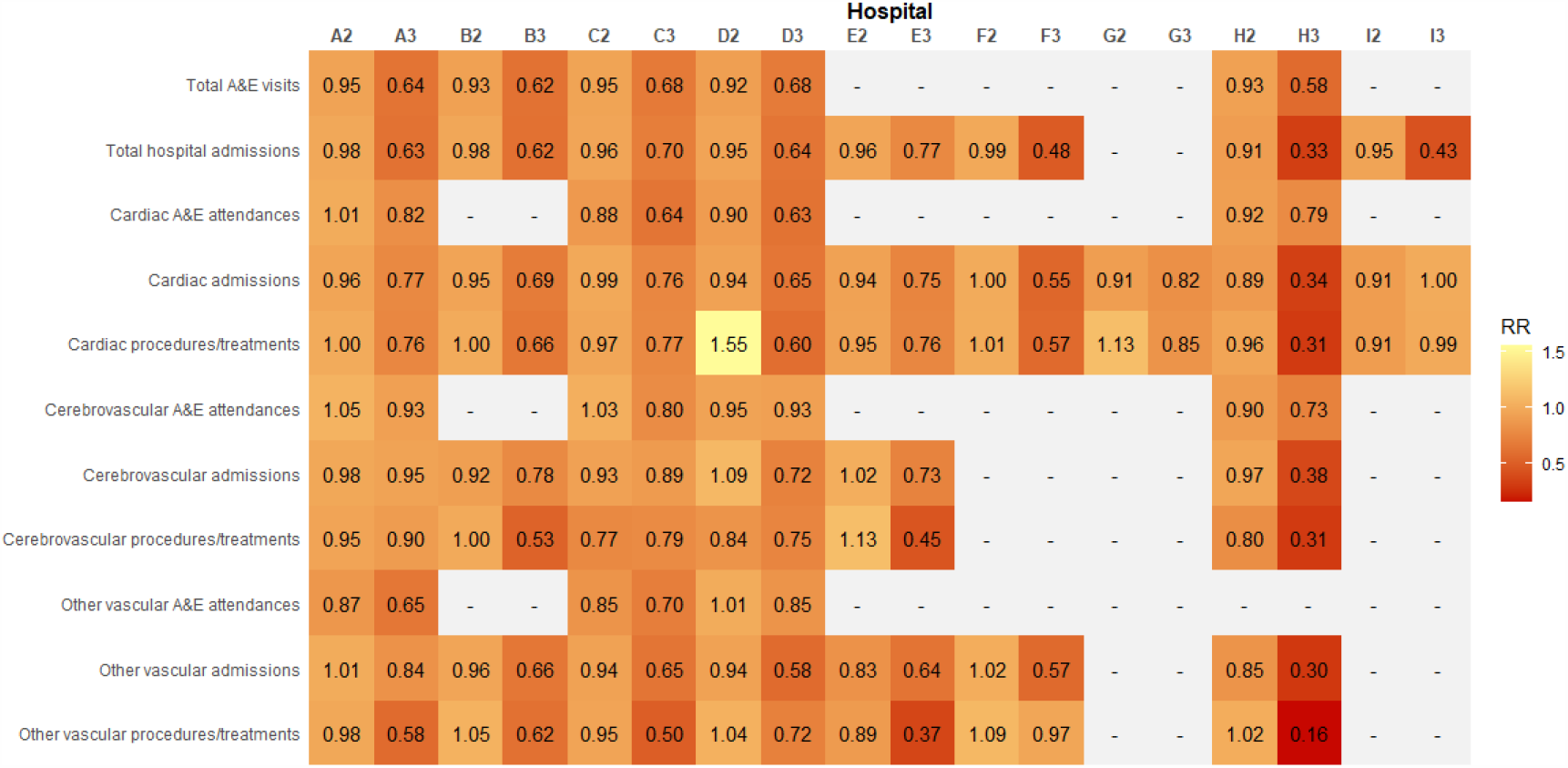
Relative change in hospital activities compared to before first case of COVID-19 across individual hospitals. Hospitals are represented as A to I with affix referring to the phase: 2 = between first case and lockdown; 3 = after lockdown.

